# Sequence analysis of travel-related SARS-CoV-2 cases in the Greater Geelong region, Australia

**DOI:** 10.1101/2020.06.08.20125898

**Authors:** Tarka Raj Bhatta, Anthony Chamings, Kwee-Chin Liew, Freya Langham, Rudi Gasser, Owen Harris, Andrew Gador-Whyte, John Stenos, Eugene Athan, Soren Alexandersen

**Author notes:** Corresponding Author; (S.A). These co-authors contributed equally (Tarka Raj Bhatta, Anthony Chamings).

## Abstract

This study reports the sequence analysis of severe acute respiratory syndrome coronavirus-2 (SARS-CoV-2) from infected individuals within the Greater Geelong region, Victoria, Australia. All but one individual had recently returned from travelling abroad, and all had clinical signs consistent with SARS-CoV-2 infection. SARS-CoV-2 belonging to three lineages were detected and represent separate introductions of the virus into the region. Sequence data were consistent with the recent travel history of each case. Full virus genome sequencing can play an important role in supporting local epidemiological tracing and monitoring for community transmission. Quality of the SARS-CoV-2 sequences obtained was highly dependent on appropriate sample collection and handling.

## Introduction

In late December 2019 a novel betacoronavirus, subsequently named severe acute respiratory syndrome coronavirus-2 (SARS-CoV-2), first appeared in Wuhan, Hubei Province, China ^1,2^. The virus could be readily transmitted from human to human by respiratory droplets and rapidly spread worldwide ^3,4^. On 25^th^ January 2020, the Australian Department of Health reported the first confirmed case of SARS-CoV-2 infection in Melbourne, Australia in a man returning to Australia from Wuhan on the 19^th^ of January 2020 ^5^. On the 7^th^ of March 2020, the city of Greater Geelong, the second largest population centre in the Australian state of Victoria, confirmed its first case of SARS-CoV-2 infection, the 20^th^ in Victoria, in a traveller returning from the USA ^6^. On the 11^th^ of March 2020, when more than 118,000 cases from 114 countries around the world had been reported, the World Health Organization (WHO) declared SARS-CoV-2 infection a global pandemic ^7,8^. Between the 15^th^ and 16^th^ of March 2020, Australia implemented travel restrictions including banning non-residents from entering the country and for all returning resident travellers to self-isolate for 2 weeks, as well as domestic movement restrictions and social distancing measures including closing of social venues and restriction or limit on the number of people gathering in one location ^9,10^.

The regional health network in the Greater Geelong region commenced SARS-CoV-2 sample collection and testing in the last week of January 2020. In addition to the state reference laboratory, local laboratories mobilised to setup diagnostic PCR capacity. We describe here next generation sequencing (NGS) and analysis of SARS-CoV-2 sequences from SARS-CoV-2 positive samples collected from 7^th^ March to the 14^th^ of April 2020 to understand the molecular epidemiology of the outbreak in the Greater Geelong region. We briefly describe how sequence analysis can support local epidemiological investigations. We also show how proper sample collection and handling influence overall sequence quality.

## Results

### Virus sequencing of SARS-CoV-2 positive samples

Ampliseq NGS for SARS-CoV-2 was attempted on the 13 positive samples and the one negative sample as a control (Table 1). All samples were taken through the Ampliseq process, however, two samples (GC-17 and GC-22, Table 1) failed to amplify sufficiently during the Ampliseq panel PCR and were not processed further. These were also the highest Ct samples for which sequencing was attempted with Ct values of 33 and 34 respectively. The remaining 11 positive samples (from 7 individuals) and 1 negative sample (GC-28, Table 1) were sequenced and generated a total of approximately 54 million reads with an average of 4.9 million reads per sample (1.1 million - 14.3 million). The negative sample was confirmed negative for SARS-CoV-2 reads by NGS. The coverage and quality of the obtained reads of the 11 positive samples varied and correlated somewhat with the virus load as estimated by the RT-PCR, except for one sample (GC-25, Table 1) which despite a high virus load generated short reads and poorer sequence. Overall, we were able to assemble near full length SARS-CoV-2 genome consensus sequences of 6 samples (designated VIC-CBA1 to VIC-CBA6, Table 1) while 5 samples (GC-12, GC-20, GC-21, GC-25 and GC-51, Table 1) only generated partial virus coverage although, except for GC-25, having reads of high quality. The near complete genomes came from samples with a RT-PCR Ct of 31 or lower, while the sequences with partial coverage, except for GC-25, all came from samples with a Ct of 31 or higher.

**Table 1.** Basic details of 14 samples included in this study. The results of the SARS-COV-2 RT-PCR test (Ct value), the sequence names and GISAID accession numbers are shown. Samples collected from the same individual at different dates are highlighted with same colour.

| **GCEID Sample ID** | **Travel History** | **Clinical Symptoms** | **Occupation** | **Sampling date** | **SARS-COV-2 RT-PCR Test (Ct Value)** | **Shortened Sequence Name** | **GISAID Accession** |
| --- | --- | --- | --- | --- | --- | --- | --- |
| GC-28 | Hong Kong | Fever, cough, sore throat, body pains, chest pain, non-productive cough | Non health care worker | 28/01/2020 | Not Detected | - | - |
| GC-26 | US | Sore throat, dry cough | Non health care worker | 7/03/2020 | Detected (21) | VIC-CBA4 (CBA4_03_07) | EPI_ISL_430064 |
| GC-13 | Europe | Body aches, headaches, dry cough, shortness of breath | Health care worker | 23/03/2020 | Detected (29) | VIC-CBA1 (CBA1_03_23) | EPI_ISL_420855 |
| GC-11 | UK | Cold, sinusitis | Health care worker | 24/03/2020 | Detected (19) | VIC-CBA2 (CBA2_03_24) | EPI_ISL_420876 |
| GC-12 | No travel history | Sore throat, rigor, fever | Health care worker | 24/03/2020 | Detected (31) | - | Partial sequence |
| GC-14 | UK | Unspecified | Health care worker | 28/03/2020 | Detected (18) | VIC-CBA3 (CBA3_03_28) | EPI_ISL_420877 |
| GC-17 | Canada/US | Sinusitis gradually got worse | Non health care worker | 1/04/2020 | Detected (33) | - | - |
| GC-20 | UK | Cold, sinusitis | Health care worker | 2/04/2020 | Detected (31) | - | Partial sequence |
| GC-21 | UK | Shortness of breath, cough, rhinorrhoea and sore throat | Non health care worker | 3/04/2020 | Detected (31) | - | Partial sequence |
| GC-22 | UK | Cold, sinusitis | Health care worker | 3/04/2020 | Detected (34) | - | - |
| GC-24 | UK | Cold, sinusitis | Health care worker | 7/04/2020 | Detected (31) | VIC-CBA6 (CBA6_04_07) | EPI_ISL_430066 |
| GC-23 | UK | Asymptomatic | Health care worker | 8/04/2020 | Detected (31) | VIC-CBA5 (CBA5_04_08) | EPI_ISL_430065 |
| GC-25 | UK | Sore throat, hoarse voice | Non health care worker | 10/04/2020 | Detected (19) | - | Partial sequence |
| GC-51 | UK | Asymptomatic | Health care worker | 14/04/2020 | Detected (31) | - | Partial sequence |

The six near full length sequences, VIC-CBA1 to VIC-CBA6, were deposited in GISAID under accession numbers EPI_ISL_420855, EPI_ISL_420876, EPI_ISL_420877 and EPI_ISL_430064 to EPI_ISL_430066 and were obtained from swabs taken from four individuals. Two individuals had two separate swabs taken 11-15 days apart which generated near complete sequence. Interestingly, for one of these multiple sampled individuals from which two near complete virus genomes were obtained (GC-11 and GC-24, Table 1), another two intervening samples produced only partial sequence (GC-20) or did not amplify in the Ampliseq PCR (GC-22) (Table 1). For the other individual for which multiple samples were sequenced, GC-14 and GC-23 sampled 11 days apart, contributed good sequences while a later sample (GC-51, Table 1) obtained 17 days after the first sample (GC-14), only provided partial sequence (Table 1). In total 6 near full length SARS-CoV-2 sequences were obtained and used for further analysis.

### Phylogenetic and Network analysis

The 6 near full length consensus sequences of SARS-CoV-2 were compared to related sequences available in GISAID as of April 23, 2020. Fourteen representative closely related sequences, as well as the two earliest SARS-CoV-2 virus sequences from Wuhan, China (WH01_12_26; GISAID accession number EPI_ISL_406798 and WH04_01_05; GISAID accession number EPI_ISL_406801) were included in this analysis (Table S1). A neighbour-joining tree was generated based on nucleotide differences and rooted on the WH01_12_26 sequence as the earliest virus sequence available (Figure 1, for details of abbreviated sequence names see Table S1). The same sequences were analysed by maximum parsimony network analysis ^11,12^ and arranged by date of sampling (Figure 2). The first SARS-CoV2 positive sample from Greater Geelong (sampled on 7^th^ March 2020), designated CBA4_03_07, clustered together with sequences reported from the USA (UW3895_03_27; shown as a representative sequence) and other sequences from Victoria (VIC08_03_15) in both analyses. Interestingly, this individual had returned from travelling in the USA (Table 1). This sequence belonged to a lineage more similar to WH04_01_05 than WH01_12_26 and has been designated as lineage A.1 in Pangolin COVID-19 Lineage Assigner ^13^ (https://pangolin.cog-uk.io/) (Figure 1 and 2). The other near full length virus sequences belonged to lineages more related to the WH01_12_26 virus, but which have a characteristic aspartic acid to glycine (D-G) amino acid substitution within the spike glycoprotein (S) region caused by a change in one nucleotide at position 23403 (Table 2). The second individual with full length sequence of SARS-CoV-2, designated CBA1_03_23, was closely related to virus sequences obtained around the same time from Victoria (VIC311_03_23), the USA (UW213_03_13) and Europe (Figure 1 and 2). This individual had recently returned to Australia from Europe (Table 1). Sequences CBA3_03_28 and CBA5_04_08, from the third individual but sampled 11 days apart and who also had recently returned from the UK, were closely related to CBA1_03_23 (only 2 nucleotide difference) and to virus sequences from Victoria (VIC52_03_12) (Figure 1–2). These three sequences, CBA1_03_23, CBA3_03_28 and CBA5_04_08 were more closely related to WH01_12_26 and belonged to the lineage B.1, with the characteristic 3-nucleotides substitution (GGG to AAC) at nucleotides 28881-28883 (Figure 1–2 and Table 2) in the nucleocapsid phosphoprotein (N protein) gene. These changes result in a 2 amino acid change from arginine-glycine (RG) to lysine-arginine (KR) (Table 2), and therefore a gain of an additional basic amino acid in this part of the N protein. CBA2_03_24 and CBA6_04_07, from the fourth individual sampled 14 days apart, generated identical sequences and were relatively closely related to virus sequences from England (Engl295_03_25) and Victoria (VIC196_03_19) belonging to lineage B.1.13 ^13^ (Figure 1–2). This individual reported recently returning from the UK and was tested after developing symptoms of mild upper respiratory tract infection (Table 1).

**Table 2.** Change of nucleotides and corresponding amino acids within the Greater Geelong SARS-CoV-2 sequences when compared with WH01_12_26 (GISAID accession: EPI_ISL_406798). Nucleotide changes are shown in bold. UTR: Untranslated Region, ORF: Open Reading Frame, non-structural protein (nsp), S (Region): Surface glycoprotein, N (Region): Nucleocapsid phosphoprotein, N/A: Not applicable.

| S. N | Nucleotide Position | Region | Nucleotide |  |  |  |  |  | Amino Acid Change | Amino acid type |
| --- | --- | --- | --- | --- | --- | --- | --- | --- | --- | --- |
|  |  |  | VIC-CBA 1 | VIC-CBA 2 | VIC-CBA 3 | VIC-CBA 4 | VIC-CBA 5 | VIC-CBA 6 |  |  |
| 1 | 241 | 5' UTR | **T** | **T** | **T** | C | **T** | **T** | N/A | N/A |
| 2 | 2416 | orf1a (nsp2) | C | **T** | C | C | C | **T** | NO | N/A |
| 3 | 3037 | orf1a (nsp3) | **T** | **T** | **T** | C | **T** | **T** | NO | N/A |
| 4 | 4510 | orf1a (nsp3) | G | **T** | G | G | G | **T** | NO | N/A |
| 5 | 7765 | orf1a (nsp3) | C | **T** | C | C | C | **T** | NO | N/A |
| 6 | 8782 | orf1a (nsp4) | C | C | C | **T** | C | C | NO | N/A |
| 7 | 11638 | orf1a (nsp6) | T | T | **C** | T | **C** | T | NO | N/A |
| 8 | 14408 | orf1b (nsp12) | **T** | **T** | **T** | C | **T** | **T** | P to L | Both Aliphatic |
| 9 | 17747 | orf1b (nsp13) | C | C | C | **T** | C | C | P to L | Both Aliphatic |
| 10 | 17858 | orf1b (nsp13) | A | A | A | **G** | A | A | Y to C | Aromatic to Sulphur Containing |
| 11 | 18060 | orf1b (nsp13) | C | C | C | **T** | C | C | NO | N/A |
| 12 | 20578 | orf1b (nsp15) | G | **T** | G | G | G | **T** | V to L | Both Aliphatic |
| 13 | 22326 | S | **T** | C | **T** | C | **T** | C | S to F | Hydroxylic to aromatic |
| 14 | 23403 | S | **G** | **G** | **G** | A | **G** | **G** | D to G | Acidic to aliphatic |
| 15 | 24694 | S | A | A | A | **T** | A | A | NO | N/A |
| 16 | 25563 | ORF3a | G | **T** | G | G | G | **T** | Q to H | Amidic to Basic |
| 17 | 27670 | ORF7a | G | **T** | G | G | G | **T** | V to F | Aliphatic to Aromatic |
| 18 | 28144 | ORF8 | T | T | T | **C** | T | T | L to S | Aliphatic to hydroxylic |
| 19 | 28881 | N | **A** | G | **A** | G | **A** | G | RG to KR | Basic + aliphatic to both basic |
| 20 | 28882 | N | **A** | G | **A** | G | **A** | G |  |  |
| 21 | 28883 | N | **C** | G | **C** | G | **C** | G |  |  |
| 22 | 29066 | N | A | A | A | A | **G** | A | T to A | Hydroxylic to aliphatic |
| 23 | 29708 | 3' UTR | C | C | **T** | C | **T** | C | N/A | N/A |

**Figure 1.**
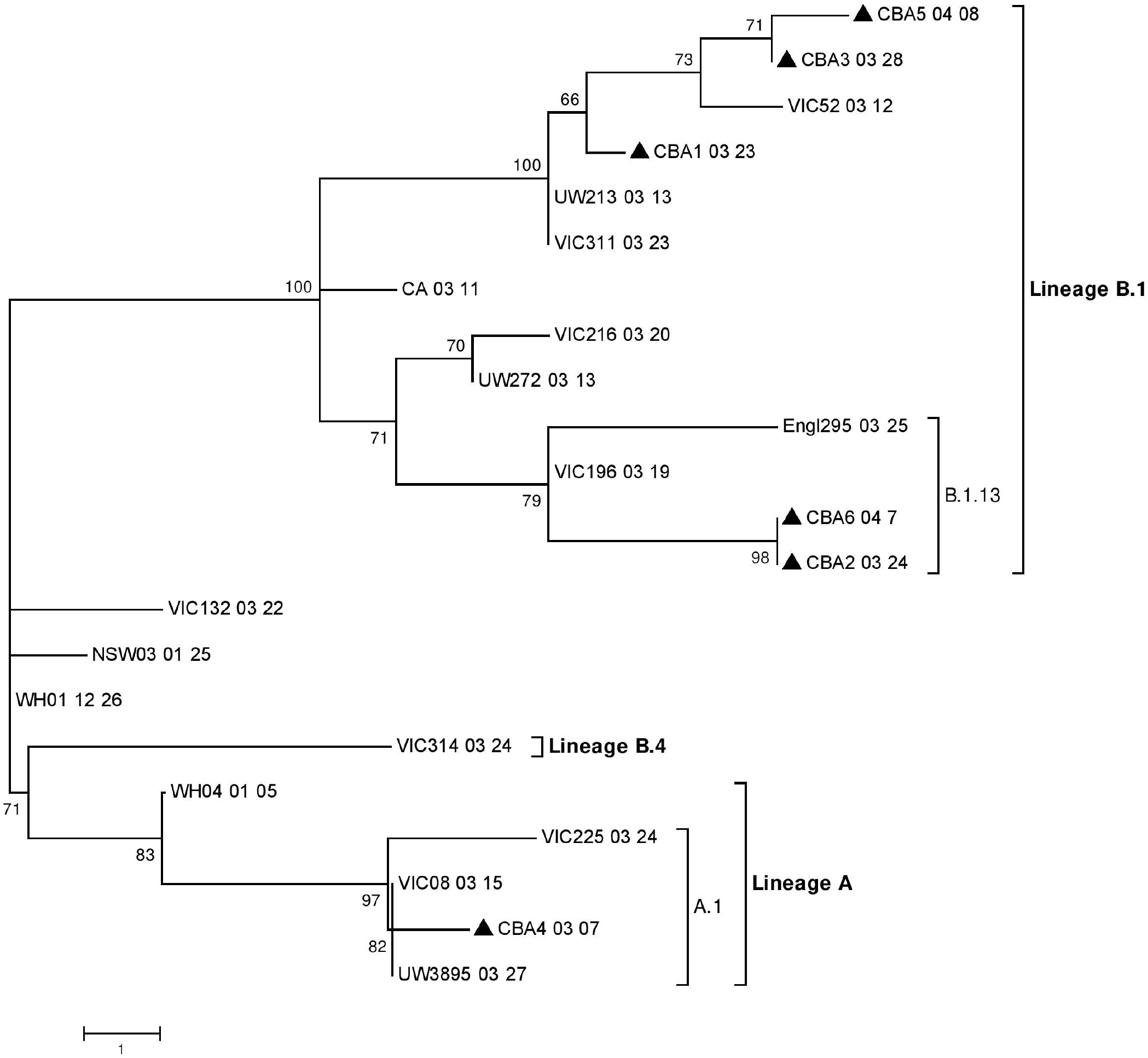
Phylogenetic analysis of the near full-length sequences of SARS-CoV-2. The nucleotide sequences were aligned and analysed using the Neighbour Joining method based on nucleotide differences and rooted on the WH01_12_26 belonging to lineage B in MEGA 7.0 ^20^ with a bootstrapping of 1000 replicates. The analysis involved 22 SARS-CoV-2 sequences (Table S1), including 6 sequences from this study. Branch lengths are scaled according to the number of nucleotide differences. The sequences from the current study have been labelled with a black triangle (▲).

**Figure 2.**
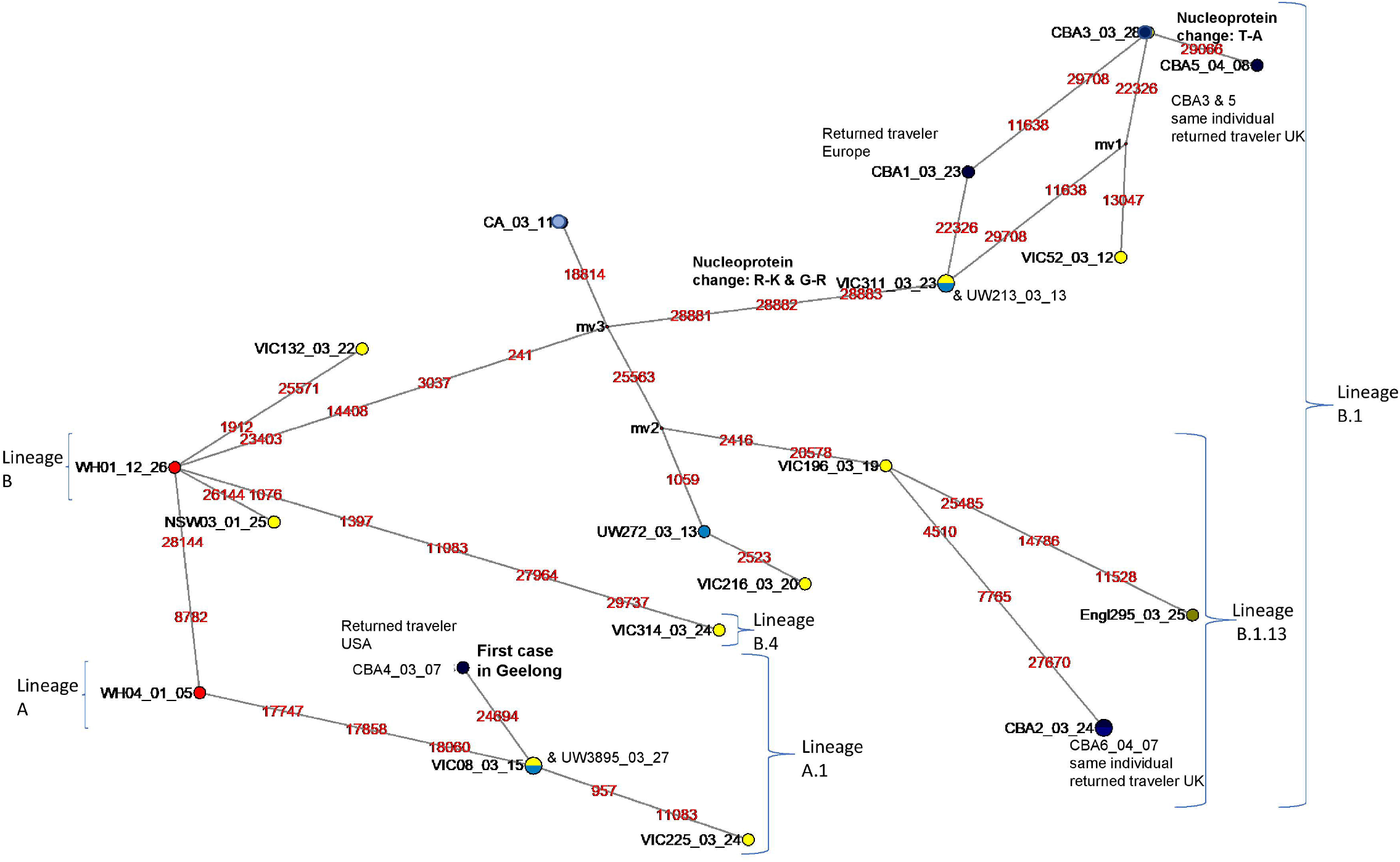
Maximum parsimony phylogenetic network analysis of the near full length sequences of SARS-CoV-2 using Network v5 ^23^ with an epsilon value of 10. The analysis involved 22 SARS-CoV-2 sequences (Table S1), including 6 sequences from this study. Each unique sequence is represented by a coloured circle showing the identity and frequency in the dataset. Branch length is proportional to the number of nucleotide differences and the position of changed nucleotide is shown in red. The larger circle labelled CBA2_03_24 also includes sequence CBA6_04_07 as these two sequences were identical. Other circles with mixed colour represent two identical sequences.

Interestingly, although sequences CBA3_03_28 and CBA5_04_08 were from samples obtained from the same individual taken 11 days apart, the later sequence, CBA5_04_08, appeared to have an additional change at nucleotide 29066 in the N protein resulting in an amino acid change from a threonine to an alanine (T-A) (see Table 2 and detailed section on amino acid changes below). A third sample (GC-51, Table 1) was obtained from this individual six days after the second sample, and seventeen days after the initial sample was collected, however only partial sequence was obtained. This partial sequence also had the AAC sequence at nucleotides 28881-28883 characteristic for this lineage, although no reads were obtained for the region at nucleotide 29066 where CBA3_03_28 and CBA5_04_08 differed (Figure 2). The sequence data from GC-51 also identified a deletion at nucleotide 15951 changing the reading frame and immediately running into a stop codon at position 15953-15955 (TAG). In addition, this sample, GC-51, also had a nucleotide change at nucleotide position 16000 (C to T), both differences not seen in any other of our sequences.

### Amino acid polymorphisms in our SARS-CoV-2 sequences

Comparison of the obtained six near full length SARS-CoV-2 sequences revealed changes at 23 nucleotide positions of which 13 changed amino acid when compared with the WH01_12_26 sequence (Table 2). While these sequences overall had 6 nucleotide changes in the ORF1a region, none of these changed the amino acid sequence. In contrast, 4 out of 5 nucleotide changes in the ORF1b (nsp12, 13 and 15) region changed amino acids (Table 2). Similarly, 2 out of 3 nucleotide changes resulted in amino acid changes in the surface glycoprotein (S) region including one mentioned above at nucleotide 23403, while each single nucleotide changes in ORF3a, ORF7a and ORF8 regions, resulted in amino acid changes in their respective proteins. The 3 nucleotides at 28881-28883 affects two codons and results in a change in two amino acids while a third change at 29066 also change the corresponding amino acid in the N protein (Table 2).

## Discussion

In this study we describe the use of NGS to complement the epidemiological and clinical investigation of SARS-CoV-2 infections detected in the Greater Geelong region, Victoria, Australia during the period from the first local case on 7^th^ March until 14^th^ April 2020. Phylogenetic and network analysis of 6 near full length sequences of SARS-CoV-2 indicated at least three or more separate introductions of the virus into the region. All cases were initially symptomatic and with a history of recent travel or working in health care, prompting their testing. The earliest introduction of SARS-CoV-2 to the Greater Geelong area on 7^th^ March 2020 (CBA4_03_07, Table 1) belonged to a lineage related to WH04_01_05 (GISAID accession number, EPI_ISL_406801), currently believed to be one of the phylogenetically earliest SARS-CoV-2 sequences and designated as lineage A.1 in Pangolin COVID-19 Lineage Assigner ^13^. This lineage was commonly detected in the United States from where this traveller had recently returned. The full length sequence (CBA1_03_23, Table 1) from the second individual and the two full length sequences (CBA3_03_28 and CBA5_04_08, Table 1) from the third individual, belonged to lineage B.1, which was commonly reported from cases in Europe and the United Kingdom ^13^. These lineages have a characteristic 3-nucleotides substitution (GGG to AAC) at nucleotides 28881-28883. These two individuals had recently travelled to Europe or the United Kingdom. The two near full length sequences of SARS-CoV-2 (CBA2_03_24 and CBA6_04_07) from the fourth individual belonged to lineage B.1.13 ^13^ which was commonly detected in the United Kingdom from where the individual had recently travelled.

Generally, SARS-CoV-2 positive samples with a Ct value of 31 or lower were found to generate near full length sequences of SARS-CoV-2 with the Ion Torrent Ampliseq method. In samples with a Ct value of 31 or higher, only partial virus genome sequences were obtained. However, sample GC-25 was an exception with a Ct value of 19. Very poor sequence data was obtained from this sample, and it was later determined that the swab from this individual had initially been placed in sterile water as opposed to an isotonic buffer, probably contributing to disruption of any coronavirus virions and degradation of the genomic RNA. In addition, we observed poor virus sequencing in two samples (GC-20 (Ct31) and GC-22 (Ct34)) taken 24 hours apart from one individual and in a sample from other individual (GC-21, Ct 31) taken during a local shortage of high quality swabs on 2-3 April 2020. A subsequent sample (GC-24, Table 1) taken from the same individual as GC-20 and GC-22, but three days later and with the high quality swab type, resulted in a sample with a Ct value of 31 and from which a near full length sequence of SARS-CoV-2 genome could be assembled (sample GC-24, sequence CBA6_04_07).

Regional virus sequencing and detailed recording of sample collection and subsequent handling has been valuable to help in understanding the local epidemic, inform ongoing SARS-CoV-2 surveillance, and explain the variations in Ct and sequence data obtained from samples. Moreover, the findings are consistent with the epidemiological and case history data indicating that these were likely introductions of the virus to the area from returning travellers. To date, most of the early cases in the state of Victoria have been introductions from returning travellers ^14^. Ongoing virus sequencing on local RT-PCR positive samples could complement epidemiological contact tracing and be a sensitive method to detect introduction of new lineages and/or early community transmission.

## Materials and Methods

### Samples

Combined nasopharyngeal and oropharyngeal swab samples were collected from individuals in the region of Greater Geelong, Victoria, Australia between the 28^th^ of January to the 14^th^ of April 2020. Testing by real time reverse-transcription PCR (RT-PCR) occurred at either Barwon Health’s Australian Rickettsial Reference Laboratory (ARRL) or at Australian Clinical Labs (ACL). Remaining sample material left over from the initial diagnostic testing was stored at −80 °C and subsequently transferred to the Geelong Centre for Emerging Infectious Diseases (GCEID) for additional RT-PCR testing and next generation sequencing (NGS). Thirteen positive samples from a total of eight individuals were identified and were included in this study. One positive individual was sampled four times and another positive individual sampled three times to monitor the progression of their infection (Table 1). Out of the 8 positive individuals, 4 were health care workers. All except one had a history of recent travel, and all had clinical symptoms consistent with SARS-CoV-2 infection (Table 1). As a negative control, we also included a SARS-CoV-2 PCR-negative swab (GC-28, Table 1).

Samples were collected for this study with approval from the Barwon Health Human Research Ethics Committee (Ref HREC 20/56), and all participants gave their informed consent for their samples and case description to be included. All methods were carried out in accordance with relevant guidelines and regulations and all experimental protocols were approved by the Barwon Health Ethics Committee.

### Nucleic acid extraction and SARS-CoV-2 RT-PCR at GCEID

Nucleic acid extraction at GCEID was carried out on all thirteen SARS-CoV-2 positive swabs and one SARS-CoV-2 negative swab (Table 1). Nucleic acid was extracted from 50 µl of swab media using the MagMAX™ Viral/Pathogen Nucleic Acid Isolation Kit (Thermofisher Scientific, Victoria, Australia) and eluted into 90 µl elution solution using a KingFisher Flex extraction robot (Thermofisher Scientific) according to the manufacturer’s instructions. The extracted nucleic acids were tested for SARS-CoV-2 using the TaqPath™ 1-Step Multiplex Master Mix without ROX (Thermofisher Scientific, Victoria, Australia) together with the TaqPath™ COVID-19 RT-PCR Kit (Thermofisher Scientific, Victoria, Australia) using 2.5 µL of extracted nucleic acids added to a final PCR volume of 12.5 µl according to the manufacturer’s instructions. This SARS-CoV-2 RT-PCR simultaneously detected three virus targets in the ORF1ab, N Gene and S Gene, and one internal extraction control target (MS2 Phage), and was run on a QuantStudio™ Flex 6 real-time thermal cycler (Applied Biosystems™) at 25 °C for 2 min, 53 °C for 10 min, 95 °C for 2 min and 40 cycles of 95 °C for 3 sec, 60°C for 30 sec

### cDNA Synthesis, NGS and Data analysis

cDNA synthesis for NGS was performed on the nucleic acids extracted from the 13 positive samples and 1 negative sample by first incubating the RNA at 70°C for 5 minutes, and then rapidly cooling on ice. SuperScript™ VILO™ Master Mix (Thermofisher Scientific, Victoria, Australia) was used as per the manufacturers’ instructions and described previously ^15^. Obtained cDNA was then amplified using the Ion Ampliseq™ Library Kit 2.0 (Thermofisher Scientific, Victoria, Australia) ^11,12^ and a commercially available SARS-CoV-2 Ampliseq panel kindly provided by Thermofisher Scientific, Victoria, Australia. This Ampliseq panel contained 237 amplicons covering the near full genome of SARS-CoV-2 and an additional 5 amplicons targeting cellular genes in two primer pools. Amplification was done following the manufacturer’s instructions for either 21, 27 or 35 cycles depending on the estimated virus load in the samples as determined by the multiplex RT-PCR. The Ion Library TaqMan™ Quantification Kit (Thermofisher Scientific, Victoria, Australia) was used for library quantification, and the libraries were run on three Ion Torrent 530 chips in an Ion S5 XL genetic sequencer (Thermofisher Scientific) at a concentration of 50pM as per the manufacturer’s protocols and as described previously ^15,16^. The sequence reads generated were mapped to a SARS-CoV-2 reference genome (NCBI GenBank accession number MN908947) ^1^ using the TMAP software included in the Torrent Suite 5.10.1 ^17^, and visualized in Integrative Genomic Viewer ^18^ (IGV 2.6.3) (Broad Institute, Cambridge, MA, USA). Near complete and partial SARS-CoV-2 genomes were aligned using Clustal-W ^19^ in MEGA 7 software ^20^. Related representative sequences from around the world were selected and downloaded on 23^rd^ April 2020 from the Global Initiative on Sharing All influenza Database (GISAID) ^21,22^ (https://www.gisaid.org/) and used for comparative phylogenetic analysis. A phylogenetic tree was generated by using the Neighbour Joining method in MEGA 7 software ^20^. Network analysis of consensus sequences was carried out in Network v5 ^23^ (Fluxus-Engineering, Clare, England) with an epsilon value of 10 as described previously ^11^. Pangolin COVID-19 Lineage Assigner ^13^ (https://pangolin.cog-uk.io/) was used on 14^th^ May 2020 to assign lineages for the SARS-CoV-2 sequences.

### Data Availability

All the sequences generated have been deposited in GISAID under accession numbers EPI_ISL_420855, EPI_ISL_420876, EPI_ISL_420877 and EPI_ISL_430064 to EPI_ISL_430066. Additional datasets analysed in the paper can be made available from the authors upon reasonable request.

## Acknowledgements

This research was funded by Deakin University, Barwon Health and CSIRO and from the National Health and Medical Research Council (NHMRC) equipment grant number GNT9000413 to S.A. We acknowledge ARRL and ACL and their staff for providing samples and for doing the initial diagnostic SARS-CoV-2 testing and in particular acknowledge Dr. Richard McCoy from ACL for his assistance in providing samples. We gratefully acknowledge Thermofisher Scientific, Victoria, Australia, for supplying the Ampliseq panel used in this study. We also acknowledge Jason Hodge, laboratory manager of the GCEID laboratory for his technical input. Finally, we gratefully acknowledge the authors and originating and submitting laboratories for the sequences and meta-data shared through GISAID’s EpiFlu™ Database ^21^ which we have used in this study (Table S1).

## Author Contributions

S.A. initiated the study and coordinated all work carried out at GCEID. A.C. and T.R.B. collected the samples from ARRL and ACL with clinical data collection coordinated by R.G., K.C.L. and F.L. O.H., A.G.W., J.S. and E.A. collated and linked the case and diagnostic test data. A.C. and T.R.B. performed the laboratory work. T.R.B., A.C. and S.A. carried out the NGS data analysis. T.R.B. drafted the initial manuscript together with S.A. and inputs from A.C. and later versions were based on input and suggestions from all. All authors contributed to the final submitted version. All authors have read and agreed to the final version of the manuscript.

## Additional Information

### Competing Interests

The authors declare no conflict of interest. The funders had no role in the design of the study, in the collection, analyses, or interpretation of data, in the writing of the manuscript, or in the decision to publish the results.

## Notes

### Competing Interest Statement

The authors have declared no competing interest.

### Author Declarations

Samples were collected for this study with approval from the Barwon Health Human Research Ethics Committee (Ref HREC 20/56).

## References

1 Wu, F. et al. A new coronavirus associated with human respiratory disease in China. Nature 579, 265–269 (2020).

2 Hui, D. S. et al. The continuing 2019-nCoV epidemic threat of novel coronaviruses to global health-the latest 2019 novel coronavirus outbreak in Wuhan, China. Int. J. Infect. Dis. 91, 264–266 (2020).

3 Chan, J. F.-W. et al. A familial cluster of pneumonia associated with the 2019 novel coronavirus indicating person-to-person transmission: a study of a family cluster. The Lancet 395, 514–523 (2020).

4 Dong, E., Du, H. & Gardner, L. An interactive web-based dashboard to track COVID-19 in real time. The Lancet infectious diseases (2020).

5 First confirmed case of novel coronavirus in Australia, <https://www.health.gov.au/ministers/the-hon-greg-hunt-mp/media/first-confirmed-case-of-novel-coronavirus-in-australia> (2020).

6 Health Alert – COVID-19 (2019 Novel Coronavirus), <https://westvicphn.com.au/about-us/latest-news/health-alert-2019-novel-coronavirus-ncov/> (2020).

7 World Health Organization, Virtual press conference on COVID-19-11 March 2020, <https://www.who.int/docs/default-source/coronaviruse/transcripts/who-audio-emergencies-coronavirus-press-conference-full-and-final-11mar2020.pdf?sfvrsn=cb432bb3_2> (2020).

8 Coronavirus disease 2019 (COVID-19) Situation Report – 51, <https://www.who.int/docs/default-source/coronaviruse/situation-reports/20200311-sitrep-51-covid-19.pdf?sfvrsn=1ba62e57_10> (2020).

9 Update on coronavirus measures, <https://www.pm.gov.au/media/update-coronavirus-measures-24-March-2020> (2020).

10 Coronavirus measures endorsed by national cabinet, < https://www.pm.gov.au/media/coronavirus-measures-endorsed-national-cabinet> (2020).

11 Alexandersen, S., Nelson, T. M., Hodge, J. & Druce, J. Evolutionary and network analysis of virus sequences from infants infected with an Australian recombinant strain of human parechovirus type 3. Sci. Rep. 7, 1–12 (2017).

12 Chamings, A. et al. Evolutionary analysis of human parechovirus type 3 and clinical outcomes of infection during the 2017–18 Australian epidemic. Sci. Rep. 9, 1–9 (2019).

13 Rambaut, A. et al. A dynamic nomenclature proposal for SARS-CoV-2 to assist genomic epidemiology. bioRxiv (2020).

14 Coronavirus (COVID-19), <https://www.dhhs.vic.gov.au/coronavirus> (2020).

15 Bhatta, T. R., Chamings, A., Vibin, J. & Alexandersen, S. Detection and characterisation of canine astrovirus, canine parvovirus and canine papillomavirus in puppies using next generation sequencing. Sci. Rep. 9, 1–10 (2019).

16 Bhatta, T. R., Chamings, A., Vibin, J., Klaassen, M. & Alexandersen, S. Detection of a Reassortant H9N2 Avian Influenza Virus with Intercontinental Gene Segments in a Resident Australian Chestnut Teal. Viruses 12, 88 (2020).

17 Caboche, S., Audebert, C., Lemoine, Y. & Hot, D. Comparison of mapping algorithms used in high-throughput sequencing: application to Ion Torrent data. BMC Genomics 15, 264 (2014).

18 Thorvaldsdóttir, H., Robinson, J. T. & Mesirov, J. P. Integrative Genomics Viewer (IGV): high-performance genomics data visualization and exploration. Briefings in bioinformatics 14, 178–192 (2013).

19 Larkin, M. et al. Clustal W and Clustal X version 2.0 Bioinformatics 23, 2947–2948 (2007).

20 Kumar, S., Stecher, G. & Tamura, K. MEGA7: molecular evolutionary genetics analysis version 7.0 for bigger datasets. Mol. Biol. Evol. 33, 1870–1874 (2016).

21 Elbe, S. & Buckland-Merrett, G. Data, disease and diplomacy: GISAID’s innovative contribution to global health. Global Challenges 1, 33–46 (2017).

22 Shu, Y. & McCauley, J. GISAID: Global initiative on sharing all influenza data–from vision to reality. Eurosurveillance 22 (2017).

23 Polzin, T. & Daneshmand, S. V. On Steiner trees and minimum spanning trees in hypergraphs. Operations Research Letters 31, 12–20 (2003).

